# Covid-19, non-Covid-19 and excess mortality rates not comparable across countries

**DOI:** 10.1101/2021.03.31.21254689

**Authors:** Gabrielle Kelly, Stefano Petti, Norman Noah

## Abstract

Evidence that more people in some countries and fewer in others are dying because of the pandemic, than is reflected by reported Covid-19 mortality rates, is derived from mortality data. Using publicly available databases, deaths attributed to Covid-19 in 2020 and all deaths for the years 2015-2020 were tabulated for 35 countries together with economic, health, demographic, and government response stringency index variables. Residual mortality rates (RMR) in 2020 were calculated as excess mortality minus reported mortality rates due to Covid-19 where excess deaths were observed deaths in 2020 minus the average for 2015-2019. Differences in RMR are differences not attributed to reported Covid-19. For about half the countries, RMR’s were negative and for half, positive. The absolute rates in some countries were double those in others. In a regression analysis, population density and proportion of female smokers were positively associated with both Covid-19 and excess mortality while the human development index and proportion of male smokers were negatively associated with both. RMR was not associated with any of the investigated variables. The results show that published data on mortality from Covid-19 cannot be directly comparable across countries. This may be due to differences in Covid-19 death reporting and in addition, the unprecedented public health measures implemented to control the pandemic may have produced either increased or reduced excess deaths due to other diseases. Further data on cause-specific mortality is required to determine the extent to which residual mortality represents non-Covid-19 deaths and to explain differences between countries.

## 1 Introduction

The pandemic of Covid-19 has caused varying excess mortality globally. The data recorded by individual countries may be inconsistent and comparison between countries unreliable. Furthermore, the effects of lockdown and other forms of prevention may also have affected mortality rates generally. It is unlike other epidemics such as influenza and HIV. With influenza and HIV there was significant mortality not officially assigned to either infection [1,2]. With Covid-19 the situation has become much more complex, since the pandemic [3] may have resulted in increased deaths from other causes such as weakened healthcare systems, fewer people seeking treatment for non-Covid diseases, less available funding for treating other diseases; or in decreased deaths from other causes, such as road accidents, mobility restrictions, air pollution, or a reduced incidence of influenza and other respiratory infections.

We observe that, precautions taken by countries to stop the spread of the disease may have led to both increases and decreases in deaths from other causes.

The pandemic is monitored by using data from sources within countries that ultimately report to the WHO. The data obtained here from the Johns Hopkins University/WHO rely on summary data supplied by countries - or, in some instances, a variety of sources.

Covid-19 deaths are defined in two main ways. The first, based on the WHO definition, uses clinically confirmed or probable Covid-19 cases (as in Belgium, Canada, France, Germany, Italy) and is not necessarily dependent on the availability of a laboratory test. The second, on the other hand, is reliant primarily on a positive laboratory test (Austria, the Netherlands, Spain, the United Kingdom) [4]. Cross-country comparisons of Covid-19 mortality rates are complex due to both different definitions of Covid-19 deaths and to under or over reporting of Covid-19 deaths. It follows that reporting of deaths due to Covid-19 can differ substantially, complicating cross-country comparisons.

Excess mortality, defined as observed minus expected mortality, makes it possible to better understand the overall impact of Covid-19 on a population [5,6]. It captures not only confirmed deaths, but also deaths attributed to other causes, which may be either increases or decreases attributable to the overall pandemic. In some countries, excess deaths during the Covid-19 pandemic are fewer than the number of reported Covid-19 deaths, while in others they are greater.

The purpose of this study is to estimate the full extent of the effects on death of the Covid-19 pandemic, the possible underestimate in the number of cases reported to the WHO, and to suggest explanations for differences between countries.

## 2 Data sources

We used data from several sources.

- Our World in Data (owid) database [7], specifically Our World in Data Covid-19 dataset: Their data on confirmed deaths due to Covid-19 comes from the COVID-19 Data Repository by the Center for Systems Science and Engineering (CSSE) at Johns Hopkins University (JHU). This is done on a country basis and the period considered here is 5/01/2020 to 2/01/2021. Other variables are also provided that are collected from a variety of sources (United Nations, World Bank, Global Burden of Disease, Blavatnik School of Government, etc.). More information is available in their codebooks. These variables include the stringency index as defined below, population size, population density, median age, aged 65 or older, aged 70 or older, GDP per capita, extreme poverty, cardiovascular death rate, diabetes prevalence, female smokers, male smokers, handwashing facilities, hospital beds per thousand, life expectancy and human development index (a composite index measuring average achievement in three basic dimensions of human development from the United Nations Development Programme: life expectancy, education, and gross national income per capita).
- The Oxford COVID-19 Government Response Tracker (OxCGRT) systematically collects information on several different common policy responses that governments have taken to respond to the pandemic on 20 indicators such as school closures and travel restrictions. The data from the 20 indicators is aggregated into an overall government response index, a number between 1 and 100 to reflect the level of government action. This stringency index for each country was also added to the data [8]. Its value changes over time and the average of the weekly values over the period of the study was used.
- We based our list of countries on the Human Mortality Database Short-term Mortality Fluctuations project (HMD) that provides detailed mortality and population data for 34 countries [9]. It was well documented and was used by the Our World in Data Covid-19 dataset on the owid website [7] where we obtained the explanatory variables. At the time of our access the HMD was the sole source of mortality data for owid. Subsequently, further countries - approximately 187 - were added in owid, and these are less well documented; data on some countries is patchy although on others it is not. The mortality data and Covid-19 data were merged to give a database for 34 (plus one below) countries: Austria, Belgium, Bulgaria, Chile, Canada, Croatia, Czechia, Denmark, Estonia, Finland, France, Germany, Greece, Hungary, Iceland, Israel, Italy, Latvia, Lithuania, Luxembourg, Netherlands, New Zealand, Norway, Poland, Portugal, South Korea, Slovenia, Slovakia, Spain, Sweden, Switzerland, Taiwan, UK and the USA. For some countries, data on observed all-cause deaths were not available for the entire period and therefore both files were merged up to the following dates. These were Greece until 12/12/2020, Italy until 5/12/2020, Slovakia until 5/12/2020, Slovenia until 26/12/2020, South Korea until 12/12/2020 and Taiwan until 3/10/2020. Since the data were sourced, we noted that updated data were now available for the entire study period. These show that the differences between the percentage of Covid deaths over all deaths, used here and updated, range between -0.001% for Taiwan and +1.710% for Slovakia; these would not greatly affect results.
- Data for Ireland was obtained from the national statistics office [10] and was available on a calendar year basis from 1/1/2020 to 31/12/2020.. Ireland was then added to the database above.

All databases were accessed in February, 2021 and that in Ireland re-accessed June 2021.

## 3 Methods

Data on the following three numbers were assessed for 35 countries:

- Observed deaths 2020,
- Average number of death 2015-2019,
- Reported Covid-19 deaths in 2020

In addition, we used available population sizes for 2020. Excess deaths were defined to be (Observed 2020 - Average 2015-2019) on a country basis. We noted Covid-19 deaths + change in deaths due to other causes (positive or negative) = Excess deaths; or Excess deaths - Covid-19 deaths = change in deaths due to other causes (positive or negative) – these we labelled as residual deaths. Residual deaths per 100,000 of population was denoted as residual mortality rate (RMR).

We included differences in reporting in ‘differences not attributed to Covid-19’. If a death was not attributed to Covid 19 but should have been, it was attributed to another cause. For deaths wrongly reported as Covid-19, we assumed ‘classified other causes of death’ as being under-reported. Differences in reporting and differences in deaths due to other causes are linked and we denoted both as being attributed to Covid-19.

Assuming a Poisson distribution for counts of total deaths in each year 2015-2020 and Covid-19 deaths respectively, the difference between excess deaths and Covid-19 deaths was tested for statistical significance i.e. the countries for which residual deaths increased or decreased significantly were found.

Regression analyses were conducted to explore the relationship between economic, demographic and health variables and residual death, Covid-19 death and excess death rates per 100,000 of the population.

We used only those explanatory variables available for all countries in these analyses and this meant omitting the variables handwashing facilities and extreme poverty listed in the first data source above. We also omitted Ireland and Taiwan as they had too few explanatory variables available.

Global spatial autocorrelation of RMR was assessed using Moran’s I as follows. The centroid of each country was found, distance between the centroids calculated and the inverse of the distance matrix used to produce a matrix of weights for Moran’s I. A correlogram was used to explore Moran’s I over different spatial lags and statistical significance determined under the assumption of randomization. [11]

All statistical analyses were performed using R. P-values are denoted by p, and values of p < 0.05 were regarded as statistically significant. The percentage of variation explained by a regression model is denoted by R^2^.

## 4 Results

The countries for which residual deaths were negative (i.e. *fewer* excess than reported Covid-19 deaths) were: Belgium, Canada, Denmark, France, Greece, Hungary, Ireland, Italy, Latvia, Norway, Slovakia, South Korea, Sweden, Taiwan, *Croatia, Luxembourg, New Zealand, Switzerland*.

Countries for which residual deaths were positive (i.e. *greater* excess than reported Covid-19 deaths) were:

Austria, Bulgaria, Chile, Czechia, Finland, Germany, Lithuania, Netherlands, Poland, Portugal, Spain, United Kingdom, United States, *Estonia, Iceland, Israel, Slovenia*.

These were significantly so apart from those italicised.

The value of RMR showed substantial variation across countries [Table]. Taiwan and Canada were at the negative and Bulgaria and Poland at the positive extreme, with rates over double the next highest RMR.

**Table:** Residual mortality rate per 100,000 population and associated p-value in () indicating significance of increase/decrease

| Country | Negative residual mortality | Country | Positive residual mortality |
| --- | --- | --- | --- |
| Taiwan | -176.3 (<0.001) | Israel | 4.6 (0.105) |
| Canada | -117.1 (<0.001) | Iceland | 6.2 (0.690) |
| Latvia | -57.9 (<0.001) | United Kingdom | 8.5 (<0.001) |
| Italy | -55.3 (<0.001) | Germany | 9.2 (<0.001) |
| Slovakia | -53.8 (<0.001) | Slovenia | 11.1 (0.177) |
| Greece | -36.1 (<0.001) | Chile | 11.3 (<0.001) |
| Sweden | -27.2 (<0.001) | Finland | 11.5 (0.013) |
| Belgium | -21.0 (<0.001) | Estonia | 13.4 (0.200) |
| Ireland | -19.5 (<0.001) | Austria | 24.7 (<0.001) |
| Norway | -13.4 (<0.001) | Lithuania | 25.1 (<0.001) |
| Hungary | -11.5 (0.008) | Netherlands | 32.2 (<0.001) |
| Croatia | -9.3 (0.150) | Portugal | 48.4 (<0.001) |
| Denmark | -9.1 (0.040) | Czechia | 48.5 (<0.001) |
| France | -7.2 (< 0.001) | United States | 49.7 (<0.001) |
| Luxembourg | -6.1 (0.620) | Spain | 50.9 (<0.001) |
| South Korea | -5.8 (<0.001) | Bulgaria | 103.0 (<0.001) |
| Switzerland | -5.4 (0.128) | Poland | 114.4 (<0.001) |
| New Zealand | -4.5 (0.274) |  |  |

The Figure shows the relationship between Covid-19 mortality rates and excess mortality rates; points above the line indicate where reported Covid-19 mortality rates were greater than excess mortality rates.

**Figure:**
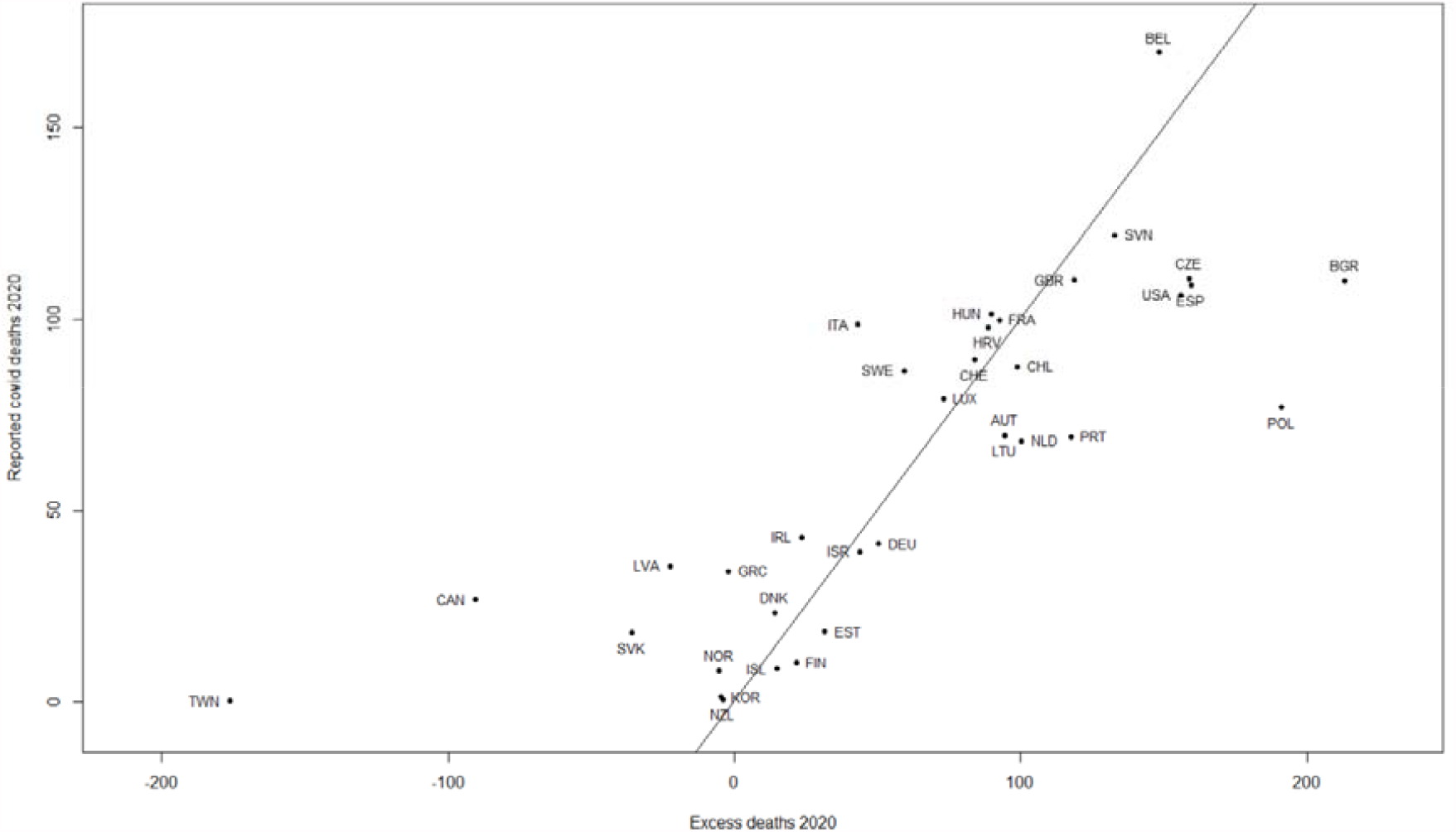
Covid deaths versus excess deaths per 100,000 of the population for 35 countries, with a line of identity. Countries are labelled by their ISO code.

The following multivariate stepwise regression analyses were carried out:

(a) Response variable: RMR. There was no association with any explanatory variable.

(b) Response variable: covid-19 mortality per 100, 000. Positive association with population density (p < 0.01), proportion of female smokers (p < 0.001), and negative association with male smokers (p < 0.01) and human development index (p=0.03). R^2^ =42%.

(c) Response: excess mortality per 100, 000. Positive association with population density (p =0.02), proportion of female smokers (p < 0.01), and negative association with male smokers (p =0.03) and human development index (p=0.04). R^2^ =34%.

Interaction terms added to the final models in (b) and (c) were not statistically significant. Model diagnostics were carried out and were satisfactory except that the Netherlands and Belgium were influential (large Cook’s distance) in both (b) and (c). Omitting these countries resulted in the same significant associations but with smaller p-values.

The correlation coefficient and 95% confidence interval (C.I.) between excess mortality and Covid-19 mortality was 0.796, (0.629,0.892), between excess mortality and RMR was 0.867 (0.750,0.931) and between Covid-19 mortality and RMR was 0.388 (0.062,0.638).

There was no evidence of spatial clustering of residual mortality, Covid 19 mortality or excess mortality using Moran’s I. For residual mortality only, the correlogram showed positive association for countries with distances in the third quartile of all distances (p=0.01).

## 5 Discussion

The results show that published data on mortality from Covid-19 cannot be directly comparable across countries. There is considerable variation in excess mortality and RMR indicating there are deaths in some countries that could be attributed to Covid-19, other than those reported. There are two factors that affect residual mortality rates (RMR) - the Covid-19 mortality rate as reported and the change in mortality rate due to other causes. If the reported Covid-19 mortality rate is accurate, or close to it, RMR would provide a way of assessing the change in deaths from other causes not attributed to Covid-19. The results show that RMR did not differ significantly from zero in eight countries: Croatia, Estonia, Iceland, Israel, Luxembourg, New Zealand, Slovenia, and Switzerland. In the remaining 27 countries 13 had a significant increase and 14 a significant decrease. There was no apparent distinguishing characteristic between countries for which the RMR was positive or negative.

Considering first countries for which the RMR was negative, Taiwan was the most extreme. The pandemic seems to have had a smaller impact in Taiwan than in most other industrialized countries, with just seven deaths in 2020. Taiwan engaged in early actions to stop the spread of the disease, including early screening of flights from mainland China and the tracking of individual cases. As discussed in [12], the experience with Ebola, MERS, and SARS meant that several Asian and African nations, including Taiwan, had systems in place to mitigate the severity of the pandemic. Other authors reported that the adjusted pneumonia and influenza mortality rates in 2020 in Taiwan were significantly lower than the 2019 rate and a deviation from the overall increasing trend since 2008 [13]. These authors concluded that the low pneumonia and influenza mortality rates in 2020 suggested that mask wearing, physical distancing, and restricting large social events may have had a positive spill over effect reducing total mortality.

The RMR in Canada was also considerably lower than in other countries. Deaths attributed to Covid-19 cannot be attributed to other causes; consequently rates of other causes of death may be reduced. Based on the WHO definition, Covid-19 is considered the first cause of death when it is thought that without the virus the patient would have not died in that moment, independently from the severity of her or his concurrent conditions. This may have happened in Canada where long-term care homes were heavily impacted by the pandemic [14].

For another 12 countries RMR was also significantly negative. Without cause-specific mortality in these countries, reasons for this cannot be ascertained definitively. Changes in behaviour may have lowered mortality from other causes too, such as traffic accidents, other accidents, other infectious diseases, as well as pneumonia and influenza. Using WHO data for six countries in the southern hemisphere: Australia, Argentina, South Africa, Paraguay, New Zealand and Chile, the total number of influenza tests fell by just 20%, while the proportion of positive tests reached record lows [14]. This indicates these countries did not have an influenza season perhaps due to social distancing and wearing of masks [13]. This seems to have been a global phenomenon as not a single case of influenza was reported in England in 2021 and a lower case rate was also reported for the U.S. [15]. These lower rates may have had a knock-on effect leading to a decrease in death rates due to flu-like illnesses. There may be some evidence that a significant decrease in travel may have caused less air pollution and thus less respiratory mortality. [16,17]

In addition, increased physical exercise in the population may have resulted in decreased deaths due to other causes [18] but there are varying reports on this. An app based study [19] reported decreased physical exercise with worldwide step count decreases. As countries impose control measures, deaths from causes such as road accidents and homicides may also have declined.

For 13 countries residual mortality was significantly positive. For these countries excess deaths were greater than those reported as Covid-19. Possible explanations include misdiagnosed Covid-19 leading to under-reporting to the WHO, whose figures may be an appreciable underestimate of the true picture. RMR’s were significantly higher in Bulgaria and Poland, double those of the next highest country. By 18 December 2020, among twenty-six European countries, Bulgaria had the highest cumulative excess mortality rate among under 65s (12.3% above the five-year average) [20]. To understand this, data on the actual causes of death is needed - it is possible that these countries could have higher death rates due to Covid-19 than those published. Wuhan where the pandemic is assumed to have originated, had a very high RMR and an 8-fold increase in pneumonia mortality in the first three months of the pandemic suggested that Covid-19 went largely underreported. Interestingly, mortality from noncommunicable diseases also increased by 20% [21].

There are two contributing factors to differences in RMR. The first is that RMR will be higher if Covid-19 mortality is under-reported and the second is that it will increase with increased mortality from other causes. The first is due to limited testing and problems in the attribution of the cause of death and this varies by country. The second may be caused by a secondary mortality due to lower or delayed access to health care, or to resources being diverted to dealing with the pandemic. Cause-specific mortality data may throw light on these two factors. In addition, as suggested in [22], social determinants of health such as jobs, income, may also be a factor in increased mortality from other causes. It was reported in the UK [23] that cardiovascular disease, went untreated or was less well treated during the pandemic. The CDC reported in late June that in the 10 weeks after the pandemic was declared a national emergency on March 13, 2020, hospital emergency department visits declined by 23% for heart attacks, 20% for strokes and 10% for uncontrolled high blood sugar in people with diabetes [24]. In the study [22] of US deaths in March and April 2020, large increases in mortality from heart disease, diabetes, and other diseases in the US were found. There is also a possibility that these increases may indicate other effects of Covid-19. Other studies in the literature report similar findings [25,26]. These changes occurred after lockdown measures were instigated.

The results of the regression analysis showing for regression analyses (b) and (c) the same significant covariates are consistent with the high correlation coefficient between excess mortality and Covid-19 mortality and their significant overlap. Analysis (a) for RMR had no significant covariates: - possibly since – residual =excess-covid and since excess and Covid mortality were similarly related to the explanatory variables, their differences cancelled these effects. Differences in RMR are complex and our covariates did not include disruptions of measures to health services or under/over-reporting. The World Health Organization (WHO) in a first indicative survey on the impact of COVID-19 on health systems based on 105 countries’ reports collected data from five regions over the period from March to June 2020. The survey illustrated that almost every country (90%) experienced disruption to its health services, with low- and middle-income countries reporting the greatest difficulties. Most countries reported that many routine and elective services were suspended, while critical care - such as cancer screening and treatment and HIV therapy – saw high-risk interruptions in low-income countries [27]. As expected by this survey, we found a high correlation coefficient between excess mortality and excess non-Covid-19 mortality i.e. RMR, suggesting that health system disruption, reduced health care demand and health care resources diverted to Covid-19 patients could be some important determinants of RMR. The correlation between Covid-19 mortality and RMR was relatively low. These results are supported by the relatively low R^2^ values in regressions (b) and (c) indicating there are other factors driving excess mortality and Covid-19 rates not considered here, and these could explain the relatively high/low correlations respectively between them and RMR. Results (b) and (c) indicate some of the same factors driving Covid-19 mortality and excess mortality. High population density makes it more difficult to control the epidemic as social distancing becomes a problem as is evident in India in 2021. The proportion of female smokers is associated with poorer countries which have fewer resources to deal with the epidemic [22]. The negative association of proportion of male smokers in analyses (b) and (c) seems counterintuitive, suggesting that active smoking is protective, at least in men. A similar finding for Covid-19 mortality and SARS-CoV-2 infection rates and male smokers has been found by other authors [28]. They suggest nicotine may modulate enzyme activity related to one of the entry doors for the Covid-19 virus within the target cells. The Netherlands was influential in the analysis because it has one of the highest population densities for these data but with high human development index and relatively low rates of male and female smokers it did not follow the general trend. Belgium was also influential because of its high rate of Covid-19 mortality. We note Belgium includes deaths in care homes that are suspected, not confirmed, as Covid-19 cases. Belgium also has a high rate of care home occupancy relative to its population [29]. This also supports the first hypothesis above that if Covid-19 is over-reported RMR will decrease.

No relationship was found with the stringency index but it must be reiterated that the value of the stringency index changes over time and an average value was used here. This is far from perfect, as countries that experienced low Covid-19 deaths were more inclined to relax lockdown measures over time.

The human development index was related to both Covid-19 and excess mortality. In Poland, for example, health expenditure was just one half the average expenditure in Europe, while per capita number of healthcare workers was at the lowest end. This healthcare system was further stressed during the pandemic, and resulted in 7% decrease in fast-track oncological cards, 15% decrease in hospitalisations for myocardial infarction, 25% decrease in stroke patients treated with mechanical thrombectomy, to cite just a few examples. Excess non-Covid-19 mortality, denoted by the second highest RMR in our list, could be a natural consequence of this situation [30].

Finally, published death rates due to Covid-19 must be interpreted with caution because there is variability between countries on how Covid-19 deaths are certified or defined [4,29,31]. In Africa, for example, cases of Covid-19 were under-reported and there is evidence the impact of Covid-19 there has been vastly underestimated [32]. A recent study [33] identifies several countries with greatly underreported Covid-19 deaths.

There was no evidence of spatial clustering of countries with respect to RMR, Covid-19 or excess mortality. This is perhaps not only because using the centroid of a country is a crude measure of location, but also because countries responded to the pandemic in a fairly independent way. More sophisticated spatial analyses confirmed this. The positive association in the correlogram at relatively large distances for RMR had no intuitive explanation from a graphical display and is probably due to factors other than spatial proximity.

There is a voluminous literature on the impact of the Covid-19 pandemic, and it was not our aim to list all relevant literature here. We have endeavoured, however, to capture the salient issues.

The paper was not meant to be comprehensive in that a study of all countries was not undertaken. This was outside the scope of the paper. A larger dataset is provided in [34]. Instead, we have provided a snapshot of well documented countries.

In conclusion, residual mortality is a useful tool to explore the indirect and direct impact of Covid-19 on mortality, as in [33]. Further data on cause-specific mortality is required to determine the extent to which residual mortality represents non-Covid-19 deaths and to explain differences between countries. Time series modelling of timing and stringency of lockdown measures on Covid-19 deaths are also necessary to assess the impact these have had in different countries.

## Data Availability

Data sources are listed in the manuscript in a separate section and all data are publicly available.

https://github.com/owid/covid-19-data/tree/master/public/data

http://data.un.org/

http://www.cso.ie

## Acknowledgements

We would like to thank the referee for many helpful comments that greatly improved the manuscript. We would also like to thank Ariel Karlinksy for comments regarding the data.

## Ethical Approval

Not applicable.

## Conflicts of interest

None.

## Funding source

None.

## Notes

### Competing Interest Statement

The authors have declared no competing interest.

### Funding Statement

No external funding was received

### Summary of Updates

Title has been changed, some data changes, some Results changes and extra material added to the Discussion.

